# Water, Sanitation and Cholera in sub-Saharan Africa

**DOI:** 10.1101/2023.02.15.23285991

**Authors:** Mustafa Sikder, Aniruddha Deshpande, Sonia T. Hegde, Espoir Bwenge Malembaka, Karin Gallandat, Robert C. Reiner, Justin Lessler, Elizabeth C. Lee, Andrew S. Azman

## Abstract

Improvements in water and sanitation should reduce cholera risk. But it is unclear which water and sanitation access are associated with cholera risk. We estimated the association between eight water and sanitation measures and annual cholera incidence access across sub-Saharan Africa (2010-2016) for data aggregated at the country and district-level. We fit random forest regression and classification models to understand how well these measures combined might be able to predict cholera incidence rates and identify high cholera incidence areas. Across spatial scales, piped or “other improved” water access was inversely associated with cholera incidence. Access to piped water, piped sanitation, and piped or “other improved” sanitation were associated with decreased district-level cholera incidence. The classification model had moderate skill in identifying high cholera incidence areas (cross-validated-AUC 0.81 95%CI 0.78–0.83) with high negative predictive values (92.5–100.0%) indicating the utility of water and sanitation measures for screening out areas that are unlikely to be cholera hotspots. While comprehensive cholera risk assessments must incorporate other data sources (e.g., historical incidence), our results suggest that water and sanitation measures could alone be useful in narrowing the geographic focus for detailed risk assessments.

**Synopsis:** We quantified the relationship between high-resolution estimates of water and sanitation access and cholera incidence and assessed the utility of water and sanitation measures in identifying high risk geographic areas in sub-Saharan Africa.

## Introduction

Access to safe water and sanitation are measured as part of global disease control initiatives and to assess progress toward the Sustainable Development Goals. Safe water and sanitation reduce the risk of water-borne diseases, improve health, and are considered fundamental human rights. The Joint Monitoring Programme (JMP), a collaboration between the World Health Organization (WHO) and United Nations Children’s Fund (UNICEF), defines improved drinking water sources as those that have the potential to deliver safe water, including piped water, boreholes, protected dug wells, protected springs, rainwater, and packaged water, and improved sanitation as those facilities designed to hygienically separate excreta from human contact.

*Vibrio cholerae*, the bacteria that causes cholera disease, is primarily transmitted through contaminated food and water. While eliminating fecal contamination of water and food by *V cholerae* should greatly reduce cholera risk, evidence documenting the impact of water, sanitation, and hygiene (WASH) on cholera in contemporary settings remains limited. This is likely due to heterogeneity in WASH intervention implementation, and the gap between access and use of safely managed infrastructure. Two systematic reviews identified low and medium-quality studies that measured the impact of short-term WASH interventions on cholera incidence; the settings, study designs, and interventions were highly variable, and the estimated reduction in cholera incidence ranged from 0 to 88% across studies and interventions.^1,2^

Ecological studies examining the association of WASH infrastructure and/or behaviors (without an explicit intervention) and cholera risk found more consistent evidence that WASH-related exposures were positively associated with cholera risk. A systematic review of individual and household-level factors found that unimproved water sources and open container water storage increased the odds of symptomatic cholera, while household water treatment and hand hygiene decreased it.^3^ Another systematic review of 51 case-control studies found that eight WASH risk factors were associated with higher odds of cholera and five out of seven WASH protective factors were associated with lower odds of cholera, although 80% of the studies were evaluated to have medium or high risk of bias.^4^ Finally, an ecological analysis found that national estimates of access to improved water sources and improved sanitation only had limited predictive value in identifying endemic cholera countries.^5^

With current global commitments to making significant progress in reducing cholera burden, including the Cholera Roadmap 2030,^6^ several countries are developing multi-year, multi-sectoral national cholera control plans. In developing these plans, countries must determine how to best utilize limited resources and seek an evidence base to help make these decisions. In this context, robust, quantitative evidence of the association between specific water and sanitation exposures and cholera risk can inform future decision making on the geographic prioritization of cholera interventions.

In our study, we leverage fine-scale estimates of water and sanitation services^7^ and suspected cholera incidence from sub-Saharan Africa^8^ to explore the association between water and sanitation infrastructure access and cholera risk at national and sub-national scales.

## Methods

### Data sources

We used previously published mean annual incidence estimates of suspected cholera (referred to throughout simply as ‘cholera’) from 2010 to 2016 in 20 km x 20 km grid cells across sub-Saharan Africa excluding Botswana, Djibouti, and Eritrea.^8^ We obtained mean annual estimates originally made at the 5km x 5km grid cell level of two sets of four mutually exclusive and collectively exhaustive indicators of access—one set for drinking water and one for sanitation.^7^ Both the drinking water indicator set (piped water on or off premises, other improved facilities, unimproved, and surface water) and sanitation indicator set (septic or sewer sanitation, other improved, unimproved, and open defecation) collectively accounts for 100% of the population in the respective geographical area. In our analysis, we grouped four putative protective measures: access to improved water, access to piped water on or off premises, access to improved (sewer or septic) sanitation, and access to piped sanitation). The remaining measures (reliance on unimproved (unprotected wells and springs) water, surface water (untreated from lakes, ponds, rivers, and streams), unimproved sanitation (flush toilets to open channels, unimproved latrines), and open defecation) were considered as putative risk factors. We obtained 1km X 1km resolution population size estimates from the WorldPop Open Population Repository for 2010 to 2016. We conducted analyses at two spatial scales: country (n=40) and second-level administrative unit level (n=4,146) with administrative boundaries based on the Database of Global Administrative Areas version 3.6.^9^

### Analysis

Our analyses included 40 countries in sub-Saharan Africa where both suspected cholera case incidence data and water and sanitation data were available. All water and sanitation measures were reported as percent of people with access to protective measures or reliance on risk measures. For the water and sanitation measures, we first calculated mean access/reliance across annual estimates from 2010 to 2016 and mean population counts across the same period; this period matched the period corresponding to the mean annual cholera incidence estimates. Then we calculated population-weighted country means for water and sanitation measures and annual cholera incidence. To quantify the association between mean annual cholera incidence and mean water and sanitation measures by country, we used univariate Quasi-Poisson regression with water and sanitation measures as linear predictors of cholera incidence and total population of the country as an offset term.

We aggregated water and sanitation and cholera measures at the second-level subnational administrative unit (*n*=4,146), hereafter “district.” We defined high cholera incidence areas as districts where >10% of the population or >100,000 people lived in a grid cell with a mean annual incidence rate >1 per 1000 cases/year, following previous work.^8^ To study the relationship between district-level mean annual cholera incidence and water and sanitation measures, we used univariate Poisson generalized estimating equations (GEE) with country as cluster variable and district population as an offset term. Estimates of risk ratios for exposures that are ordinal in nature can be challenging to interpret (e.g. risk ratio for access to improved water will include both those with ‘better’ (piped water) and ‘worse’ (unimproved water and surface water) access to water in the comparison group). Therefore, to have more interpretable relative risk estimates we arranged the water measures as piped water, piped or “other improved” water, and surface water and sanitation measures as piped sanitation, piped or “other improved” sanitation, and open defecation.

We then used random forest models to understand the potential predictive value of all water and sanitation measures combined for predicting suspected cholera incidence rates and for identifying high cholera incidence areas (e.g., identifying administrative units where incidence rates exceed a specific threshold). We conducted leave-one-district-out cross-validation to evaluate the model performance and then ran the model on the full data to understand the predictive importance of the water and sanitation measures. For the regression models, which had mean annual incidence as the dependent variable, model fit was judged by cross-validated root mean square error (cvRMSE). In classification models, meant to discriminate areas with high incidence from those without high incidence, model fit was judged by area under the cross-validated receiver operator characteristic curve (cvAUC). We performed oversampling with replacement from the minority class to address the class imbalance between high incidence areas (n=309) and areas without high incidence (n= 3,837) in each fold using previously published methods.^10^ To understand the relative importance of each water and sanitation measure within the models, we calculated the conditional permutation importance (CPI) matrix.^11^ In secondary analysis, we fit high incidence area classification models only including countries where at least one district was classified as high cholera incidence area (25 countries) and explored the performance of gradient boosting machine (GBM) models.

The data aggregation for district- and country-level estimates were completed using the *Exactextractr*,^12^ generalized estimating equations using *gee*,^13^ the random forest models were completed using the *randomForest*,^14^ variable importance were obtained using *permimp*,^11^ and GBM models were completed using *gbm*^15^ packages in R. Code and data needed to reproduce primary analyses are available at the <u>code repository</u>.

## Results

### Country-level

Country-level population weighted mean water and sanitation measures varied across the study area (Figure 1 and Table S1). ^7^ Of the four protective factors, access to improved water had the highest mean prevalence (mean: 68%, range: 34-92%) and access to piped sanitation had the lowest (10%, range: <1%-55%). Among the risk factors, reliance on open defecation had the highest (32%, range: 2-70%) and reliance on surface water had the lowest prevalence (11%, <1-30%). The 2010-2016 mean annual incidence of suspected cholera at the national-level ranged from 1.83 (95% CI 0.96-4.01) cases per 10,000 population (Sierra Leone) to 0.0003 (95% CI 0.0001-0.0005) cases per 10,000 population (Gabon).^8^

**Figure 1:**
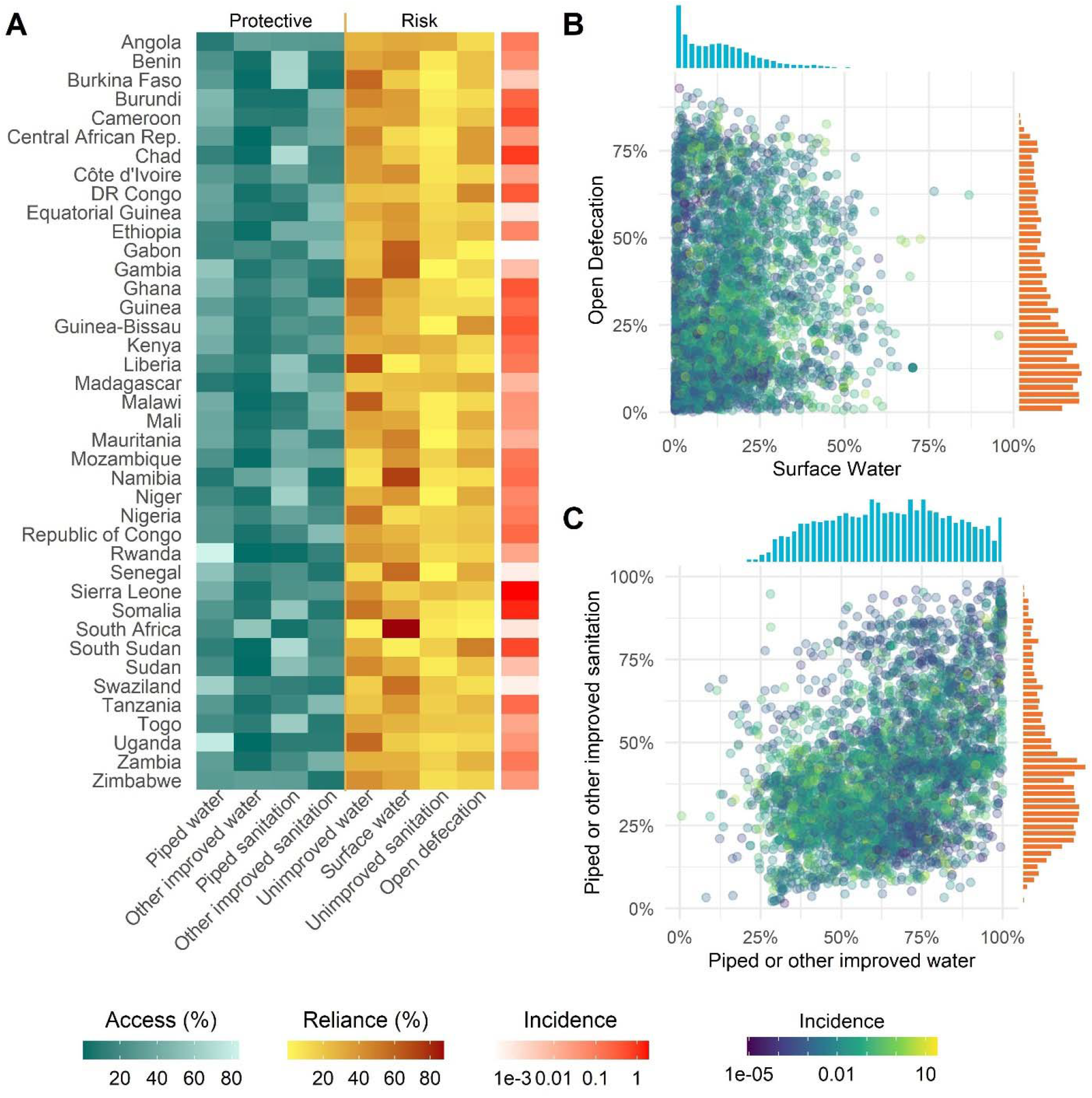
Water and sanitation measures and incidence rate of suspected cholera (2010-2016): (A) mean water and sanitation measures and log of mean annual incidence of cholera per 1,000 population by country; scatter plots with point color indicating mean cholera cases in 1,000 people (B) between reliance on surface water and open defecation (extremes) by district; (C) between improved water and improved sanitation by district. Univariate histograms of district-level measures shown along the axes of (B) and (C) are in blue and orange.

At the country-level, we found that increases in access to piped or “other improved” water were associated with a significant decrease in mean annual cholera incidence in univariate analysis. A 1% increase in piped or “other improved” water access was associated with a 7.0% (95%CI 4.0–10.0) decrease in mean annual cholera incidence within the country. The remaining putative protective factors, access to piped water (3.0%, 95%CI 0.0–6.0), piped or “other improved” sanitation (1.0%, 95%CI -1.0–4.0), and piped sanitation (3.0%, 95%CI -1.0–9.0), had point estimates consistent with being protective though they were not significantly associated with mean annual cholera incidence (Figure 2). Among the putative risk factors, none achieved statistical significance at the 0.05 level. Effect estimates for reliance on surface water and open defecation were uncertain and confidence intervals spanned the null.

**Figure 2:**
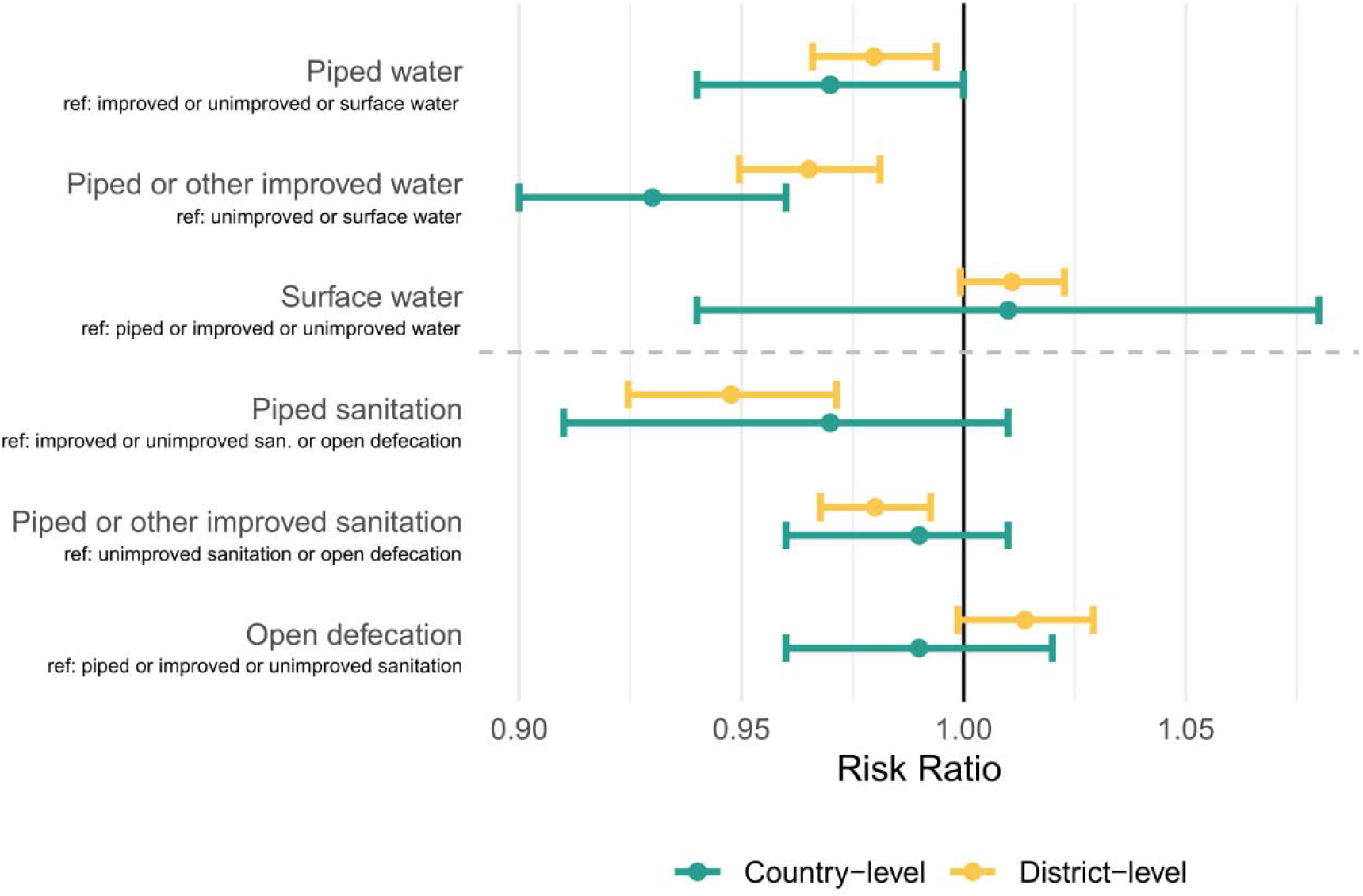
Risk ratio of water and sanitation measures from univariate models at country (Quasi-Poisson) and district scales (Poisson GEE). Reference groups (denoted ‘ref’) for each model are included to compare the risk ratios.

### District-level

When aggregating data to the district-level, results were qualitatively consistent with the country-level analyses. Increases in access to piped water, piped or “other improved” water, piped sanitation, and piped or “other improved” sanitation were associated with a significant decrease in mean annual cholera incidence (Figure 2, Table S2). For example, a 1% increase in access to piped or “other improved” water was associated with a 3.5% decrease (95%CI 1.9– 5.1) in mean annual cholera incidence and a slightly larger reduction in incidence with piped sanitation (5.2% decrease, 95%CI 2.9-7.6). The proportion of the population using surface water was associated with an increase of mean annual incidence of 1.1% (95%CI -0.1–2.3%) and open defecation was associated with an increase of mean annual incidence of 1.4% (95%CI - 0.1–2.9%), though neither was statistically significant (Figure 2, Table S2).

To better understand the potential predictive value of multiple water and sanitation measures combined for identifying high risk cholera areas, we used random forest regression models to predict mean annual cholera incidence and classification models to identify high incidence area districts. The cvRMSE of the random forest regression model was 0.92 (95% CI 0.90-0.94).

With all six water and sanitation measures, the random forest model was able to explain 37% of the observed variability in mean annual cholera incidence in districts. The cross-validation predictions were weakly correlated with the true incidence (Spearman rho = 0.60) (Figure 3). In the full data model, the most influential predictors of mean annual incidence tended to be at the extreme ends of the water and sanitation hierarchy; open defecation followed by piped sanitation, piped water, surface water, piped or “other improved” water, and piped or “other improved” sanitation (Figure S1A). Similar qualitative results were found with the GBM model (see Supplement) though the model did not fit as well (leave-one-district-out cvRMSE 1.06 95% CI 1.04-1.08) and piped water and piped sanitation were among the top three important variables in both models (Figure S1).

**Figure 3:**
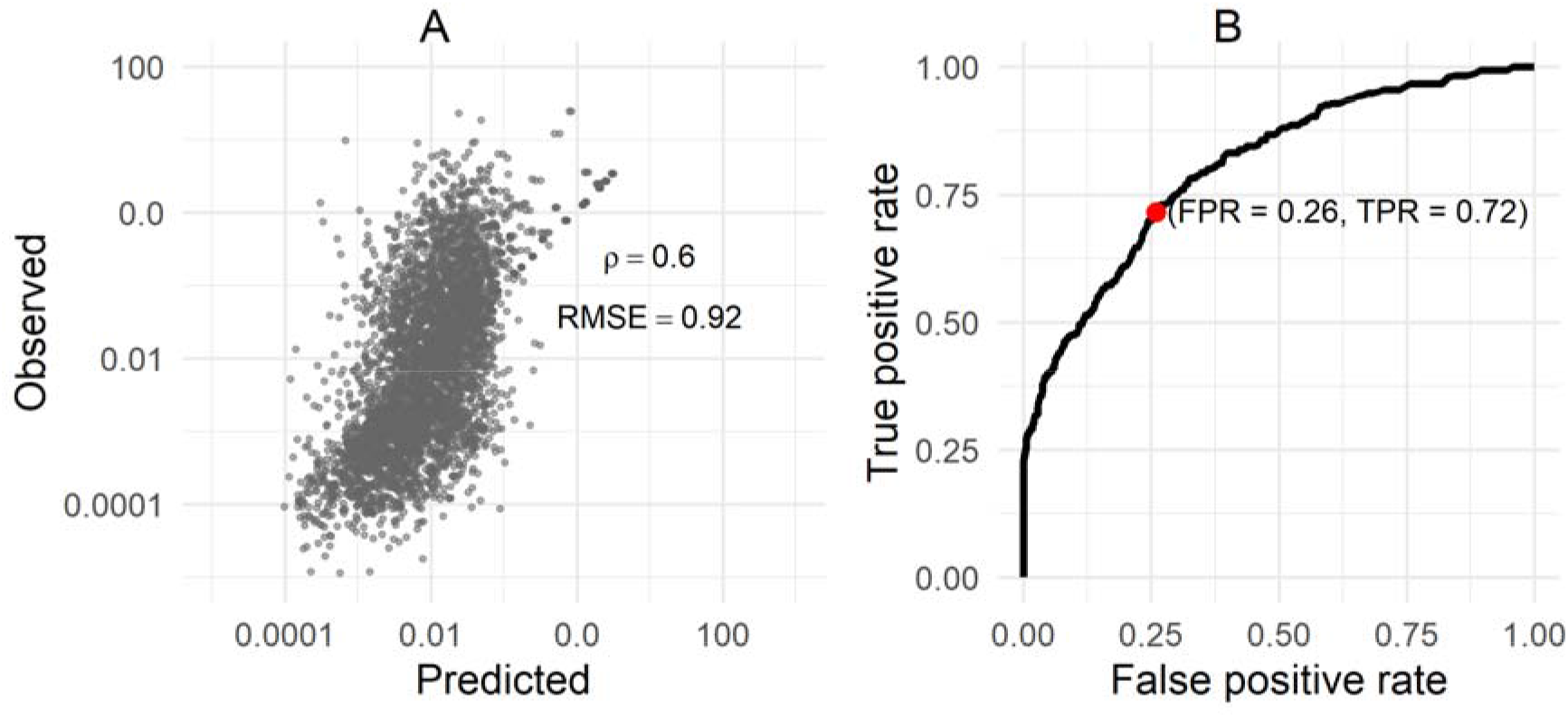
Summary of random forest models’ performance. Panel A illustrates the observed mean annual incidence versus the predicted values in cross validation. Panel B illustrates the cross-validated receiver operator characteristic curve from the random forest classification model used to identify high incidence areas. Youden cut-off point (jointly maximized sensitivity and specificity) shown as a red dot in panel B.

The random forest high incidence area classification model demonstrated moderate performance with a cvAUC of 0.81 (95% CI 0.78-0.83). At the point that maximized model sensitivity and specificity, it incorrectly classified 28.5% of true high incidence areas (1-sensitivity) and falsely classified 26.0% of districts as high incidence areas (1-specificity). While the positive predictive value of these classification models varied across cutoff thresholds (range 7.6-98.6%), the negative predictive value remained high (range 92.5-100.0%, Table S4). This high negative predictive value suggests that even in the absence of reliable cholera incidence data, water and sanitation variables, ideally in conjunction with other factors, might be useful in screening out areas that are unlikely to be at risk for having high cholera incidence.

The most influential water and sanitation measure in high incidence area classification models was surface water followed by piped sanitation, open defecation, piped water, piped or “other improved” water, and piped or “other improved” sanitation (Figure S2A).

We found similar qualitative results using the GBM model (cvAUC 0.73, 95% CI 0.70-0.76; Table S3). When we focused only on the 25 countries that had at least one high cholera incidence area, we found that performances of the models were reduced (cvAUC 0.71 (95% CI 0.68-0.74) for random forest and 0.66 (95% CI 0.62-0.69) for the GBM model; Figure S3, Table S3).

## Discussion

We investigated the relationship between water and sanitation access and suspected cholera incidence at both the national and district-level across sub-Saharan Africa. While the direction of the association between individual water and sanitation measures and cholera generally aligned with prevailing evidence and beliefs, the size and significance of these associations varied across models and geographic scale of analysis. When combining all measures together, our classification models, while far from perfect, demonstrated moderate skill in discriminating high cholera incidence areas from areas without high incidence, and may be especially useful in excluding areas as potential cholera hotspots. As most of the evidence on water and sanitation and cholera come from small scale highly local interventions, often in response to outbreaks, with outcomes measured over a short timeframe, our results help fill the evidentiary gap on the associations between multi-annual cholera incidence rates and access to water and sanitation infrastructure in sub-Saharan Africa over the same period. They highlight some of the complexities of using these water and sanitation metrics in identifying priority targets for cholera control but suggest that there is likely value in including these in risk assessment.

The variable association of water and sanitation estimates and cholera incidence between country- and district-level analyses indicates the importance of the spatial scale of analysis. Water and sanitation access can significantly vary within countries, within and between subnational units, and particularly between urban and rural areas ^16,17^, and while interventions like piped water might be implemented in whole jurisdictions, others like installation of sanitation facilities may occur on smaller scales. It is possible that more accurate and finer resolution data may have better predictive value for high cholera incidence areas, but these data are difficult to collect and rarely available when national cholera control planning is underway.

Beyond the general limitations of ecological analyses, the modeled gridded estimates of water and sanitation access and cholera incidence are imperfect given the sparse data in both data sets. Measures of access or reliance on different water and sanitation measures do not reflect behavior at the individual- or population-level, which directly influences disease risk; access is not a measure of usage and when used in analyses circumvents a step in the casual pathway leading to biased associations in either direction.^18^ Behavior data across geographies and at such large scales, however, is yet unavailable. Further, the cholera incidence estimates were based primarily on data of reported suspected, not laboratory confirmed, cases. Among suspected cases, there are almost certainly those with diarrhea caused by other pathogens than *V. cholerae* O1/O139; ^19^ similarly, under-reporting can lower the actual incidence. Additionally, these estimates represented an average over time, which may have attenuated the apparent associations between measures. Previous work has suggested that incomplete and erroneous reporting in water quality can produce misleading results ^20^, which would then propagate to modeled estimates. Finally, our analysis measures cholera risk only through mean annual incidence, but metrics like outbreak size and frequency ^21^ may better capture the relationship between water and sanitation access and cholera transmission efficiency.

Our analysis highlights the relationships between water and sanitation measures and suspected cholera incidences and the potential predictive power of these measures in estimating incidence and identifying high cholera incidence areas. Water and sanitation measures alone cannot explain the variability in cholera incidence seen throughout sub-Saharan Africa, but our analyses do suggest that they contain important information for helping to distinguish areas of high and low cholera risk. While not demonstrated in these analyses, using these measures in conjunction with data on previous incidence of cholera may help improve future guidance on assessing cholera risk. Future work to collect more precise and health-relevant metrics on water and sanitation (e.g., water quality tests, service quality indicators and other metrics following revised JMP standards on safely managed services ^22^), data on water and sanitation related behaviors, and data on confirmed, rather than suspected cholera, can help refine our understanding of these important relationships.

## Data Availability

All data produced are available online at https://www.healthdata.org/

## References

1. Taylor, D. L., Kahawita, T. M., Cairncross, S. & Ensink, J. H. J. The impact of water, sanitation and hygiene interventions to control cholera: A systematic review. PLoS ONE vol. 10 e0135676 (2015).

2. Yates, T., Vujcic, J. A., Joseph, M. L., Gallandat, K. & Lantagne, D. Efficacy and effectiveness of water, sanitation, and hygiene interventions in emergencies in low- and middle-income countries: a systematic review. Waterlines 37, 31–65 (2018).

3. Richterman, A., Sainvilien, D. R., Eberly, L. & Ivers, L. C. Individual and Household Risk Factors for Symptomatic Cholera Infection: A Systematic Review and Meta-analysis. J Infect Dis 218, S154–S164 (2018).

4. Wolfe, M., Kaur, M., Yates, T., Woodin, M. & Lantagne, D. A Systematic Review and Meta-Analysis of the Association between Water, Sanitation, and Hygiene Exposures and Cholera in Case–Control Studies. The American Journal of Tropical Medicine and Hygiene 99, 534–545 (2018).

5. Nygren, B. L., Blackstock, A. J. & Mintz, E. D. Cholera at the Crossroads: The Association Between Endemic Cholera and National Access to Improved Water Sources and Sanitation. The American Journal of Tropical Medicine and Hygiene 91, 1023–1028 (2014).

6. Global Task Force on Cholera Control. Ending Cholera — A Global Roadmap to 2030. https://www.who.int/cholera/publications/global-roadmap/en/ (2017).

7. Deshpande, A. et al. Mapping geographical inequalities in access to drinking water and sanitation facilities in low-income and middle-income countries, 2000–17. The Lancet Global Health 8, e1162–e1185 (2020).

8. Lessler, J. et al. Mapping the burden of cholera in sub-Saharan Africa and implications for control: an analysis of data across geographical scales. The Lancet 391, 1908–1915 (2018).

9. GADM. Global Administrative Areas. https://gadm.org/about.html (2020).

10. Lunardon, N., Menardi, G. & Torelli, N. ROSE: a Package for Binary Imbalanced Learning. The R Journal 6, 79 (2014).

11. Debeer, D., Hothorn, T. & Strobl, C. permimp: Conditional Permutation Importance. R package version. (2021).

12. Baston, D. exactextractr: Fast extraction from raster datasets using polygons. vol. R package Version 0.1.1 (2019).

13. Carey, V. J., Lumley, T. & Ripley, B. D. gee: Generalized estimation equation solver. (2019).

14. Liaw, A. & Wiener, M. Classification and Regression by randomForest. R News 2, 18–22 (2002).

15. Greenwell, B., Boehmke, B., Cunningham, J. & Developers (https://github.com/gbm-developers), G. B. M. gbm: Generalized Boosted Regression Models. (2020).

16. Fuller, J. A., Westphal, J. A., Kenney, B. & Eisenberg, J. N. S. The joint effects of water and sanitation on diarrhoeal disease: A multicountry analysis of the demographic and health surveys. Tropical Medicine and International Health 20, 284–292 (2015).

17. Roche, R., Bain, R. & Cumming, O. A long way to go -Estimates of combined water, sanitation and hygiene coverage for 25 sub-Saharan African countries. PLoS ONE 12, e0171783 (2017).

18. Lopez, V. K. et al. Determinants of Latrine Use Behavior: The Psychosocial Proxies of Individual-Level Defecation Practices in Rural Coastal Ecuador. Am J Trop Med Hyg 100, 733–741 (2019).

19. Wiens, K. E. et al. Towards estimating true cholera burden: a systematic review and metaanalysis of Vibrio cholerae positivity. 2022.10.05.22280736 Preprint at https://doi.org/10.1101/2022.10.05.22280736 (2022).

20. Sikder, M., Naumova, E. N., Ogudipe, A. O., Gomez, M. & Lantagne, D. Fecal indicator bacteria data to characterize drinking water quality in low-resource settings: Summary of current practices and recommendations for improving validity. International Journal of Environmental Research and Public Health 18, 1–19 (2021).

21. Zheng, Q. et al. Cholera outbreaks in sub-Saharan Africa during 2010-2019: a descriptive analysis. Int J Infect Dis 122, 215–221 (2022).

22. WHO/UNICEF. WASH in the 2030 Agenda. https://data.unicef.org/resources/wash-2030-agenda/ (2017).

